# COVID-19 transmission dynamics in South Korea prior to vaccine distribution

**DOI:** 10.1101/2024.05.17.24307538

**Authors:** Jiyeon Suh, Marta Galanti, Teresa K. Yamana, Matteo Perini, Roselyn Kaondera-Shava, Jeffrey Shaman

## Abstract

In early 2020, South Korea experienced a large coronavirus disease 2019 (COVID-19) outbreak. However, despite its proximity to China, where the virus had emerged, and the high population density of the Seoul metropolitan area, a major international hub, South Korea effectively contained the spread of COVID-19 using non-pharmaceutical interventions until vaccine distribution in 2021. Here, we built a metapopulation model with a susceptible-exposed-infectious-quarantined-recovered (SEIQR) structure and combined it with the ensemble adjustment Kalman filter to infer the transmission dynamics of COVID-19 in South Korea from February 2020 until vaccine deployment. Over the study period, the fraction of documented infections (ascertainment rate) was found to increase from 0.50 (95% credible interval (CI): 0.26—0.77) to 0.62 (95% CI: 0.39—0.86). The cumulative number of total infections, including both documented cases and undocumented infections, was less than 1% of the South Korean population at the end of the simulation period, indicating that the majority of people had yet to be infected when vaccine administration began. These findings enhance understanding of the COVID-19 outbreak in South Korea and highlight the importance of preparedness and response in managing global pandemics.

## Introduction

The coronavirus disease 2019 (COVID-19), caused by severe acute respiratory syndrome coronavirus 2 (SARS-CoV-2) emerged in Wuhan, China, in late 2019 and rapidly spread across the world, escalating into a global health crisis. In March 2020, the World Health Organization (WHO) declared a pandemic; over the next year, COVID-19 overwhelmed healthcare systems, disrupted economies, and caused millions of deaths.

South Korea was one of the first countries outside China to face a major COVID-19 outbreak during early 2020. The Korea Disease Control and Prevention Agency (KDCA) recorded its initial case on January 20, 2020, and sporadic outbreaks linked to imported cases continued in the following weeks. The first major outbreak, began on February 18, 2020, in the city of Daegu, with the confirmation of the 31st case linked to a religious gathering and an additional 7,000 cases over the following three weeks. This sudden surge significantly strained South Korea’s public health infrastructure; however, building on its experience with the Middle East respiratory syndrome coronavirus (MERS-CoV) outbreak in 2015, which resulted in nearly 186 confirmed cases and 38 deaths [1], South Korea had a strategy to address the challenges posed by COVID-19 [2-4]. The government swiftly implemented non-pharmaceutical measures such as scaled-up testing, isolating confirmed cases, and contact tracing, effectively flattening the infection curve and curtailing further spread [5]. Despite facing continual sporadic outbreaks and two significant waves centered around the Seoul metropolitan area during 2020, South Korea maintained its control strategies along with social distancing protocols without enforcing a complete lockdown, and kept per capita rates of cases, hospitalizations, and deaths low [6-8].

Modeling of COVID-19 can provide insight into infectious disease outbreaks and inform public health response and policy decisions. For the first major outbreak in Daegu, a deterministic model was previously utilized to estimate the basic reproduction number and predict future infections [9]. Public health interventions, such as quarantine, social distancing, and school closure were evaluated focusing on the entire country. Some studies segmented time horizons according to intervention policy changes to estimate piecewise transmission rates using deterministic models [10-13]. One study integrated the spatial structure of infection spread by dividing the country into two regions: Daegu and North Gyeongsang and the rest of South Korea [14], and other models have been used to generate forecasts and inform policymaking; however, most of these approaches did not represent the spatial structure of spread or consider unreported infections [15].

Studies on COVID-19 outbreaks in China and the US, using metapopulation models, have highlighted a substantial number of undocumented infections [16, 17]. Using a probabilistic model, the number of undetected infections in South Korea was estimated to range from 10,400 to 139,900 by February 2, 2021 [18]. From this range, another study derived that, on average, half of total infections were not reported [19], an estimate that is lower than the numbers inferred for China and the US. These undetected numbers may include asymptomatic or mildly symptomatic infections who do not seek testing or medical care, but still are infectious and contribute to the ongoing outbreak [16, 20]. Failing to account for these cases can underestimate the true infection rate and reproductive number, impacting the accuracy of forecasts and the modeling of intervention effectiveness.

In this study, we developed a metapopulation model coupled with Bayesian inference to analyze the transmission dynamics of COVID-19 in South Korea until the introduction of vaccines. The model is stratified into 17 si/do (city/province) levels, integrating mobility data and daily confirmed case data. Using this model-inference system, we estimated the transmission rate and ascertainment rate at the end of major outbreak waves, along with the reproduction number and the number of undocumented cases.

## Methods

### Data

Two datasets were used to initialize the metapopulation model. The first is demographic data obtained from the Statistics Korea (KOSTAT) [21]; here, we used the 2020 population census by si/do (city/province) level administrative districts in South Korea. The numbers in this dataset represent the number of residents, including both local and foreign, living in an area for that year. The second is mobility data sourced from the Korea Transportation Database (KTDB) [22], which provides annual data obtained from national transport surveys and categorized into seven purposes: going to work, going to school, business, shopping, returning home, leisure, and others. Each category has a corresponding origin and destination matrix, with its components describing the average number of daily movements from the origin to the destination. For our model, the returning home matrix for 2019 was utilized, as it represents the volume of people coming back home after going out for any reason for the year immediately prior to the COVID-19 pandemic.

To incorporate changes in mobility over time, we employed weekly updated regional movement data [23] derived from mobile phone usage of SK Telecom, which had a market share of approximately 41% in South Korea during 2021. These data provide daily intra- and inter-regional movement but at a finer administrative district (gu) level than the structure of our model. For example, Seoul consists of 25 gus. In these data, movements within a particular district, e.g. Gangnam-gu, were counted as intra-regional movements and movements from Gangnam-gu to other districts were counted as inter-regional movements. However, to be of use in our model, these inter-regional movements should be distinguished by destination. If the destination is one of the districts in Seoul, it is considered a movement within Seoul; if the destination is some district in a city other than Seoul, it is considered a movement to another si/do subpopulation. Owing to the absence of specific destination information, we aggregated both the intra- and inter-regional movement data from all 25 districts of Seoul, and this aggregation was used to adjust the baseline mobility of people moving from Seoul to other regions during the day. The same approach was applied to rescale mobility patterns for the other locations.

To calibrate the model, we used daily confirmed cases by si/do from February 8, 2020 to February 25, 2021, as reported by the Korea Disease Control and Prevention Agency (KDCA) [24]. These case numbers represent both local and imported cases.

### Metapopulation model

We built a metapopulation model with a susceptible-exposed-infectious-quarantine-recovered (SEIQR) structure to simulate COVID-19 transmission in South Korea. The model is stratified into 17 si/do levels to describe inter-regional movements. Its structure was informed by earlier COVID-19 models specific to South Korea [9, 13], as well as metapopulation models used for China and the US [16, 25].

In general, people leave their homes in the morning to go to work or school and return to their neighborhoods and homes in the evening. The model mirrors this daily pattern by dividing the entire population into 17×17 subpopulations based on origin (residence) and destination and formulating the transmission differently during the day and at night. Concretely, people who live in region *j* and move to region *i* during the day (*N*_*ij*_) mix with people in region *i* during the day and can spread infection among people in region *i*. In the evening, these people return to their residential area *j*, where they mix with people in that area and can spread infection. The time step sizes for day and night were adjusted to 1/3 and 2/3 of a day in accordance with previous modeling work [25], which showed that similar results could be obtained when the length of day and night is equal.

Each subpopulation was then compartmentalized according to its epidemiological stage as shown in Figure 1 and equations (1)—(6). Susceptible (*S*_*ij*_) individuals are infected at a rate λ _*ij*_ through contact with infectious individuals and become latent (*E*_*ij*_). The rate λ _*ij*_, known as the force of infection, is the per capita rate at which susceptible individuals become infected through contact with infectious individuals, and is proportional to the transmission rate and proportion of infectious individuals at time *t*. In this model, *λ*_*ij*_ was defined as shown in equation (7), with the transmission rate (*β* _*i*_) specified for each region, allowing for different mixing patterns during the day and night with infectious individuals. After an average of *Z* days, the latent period, individuals become infectious and bifurcate into cases that are reported (i.e. at some point tested and quarantined) with probability *α* and unreported with probability 1 − *α*. During the day, it is assumed that both reported infectious individuals 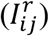 and unreported infectious individuals 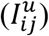 move to other regions for work or school, but for 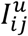 a reduced infectivity, scaled by μ, is assumed. The daytime population size of region *i* is computed according to equation (8). Note that reported infectious individuals are assumed to move normally until testing and quarantine.

**Figure 1.**
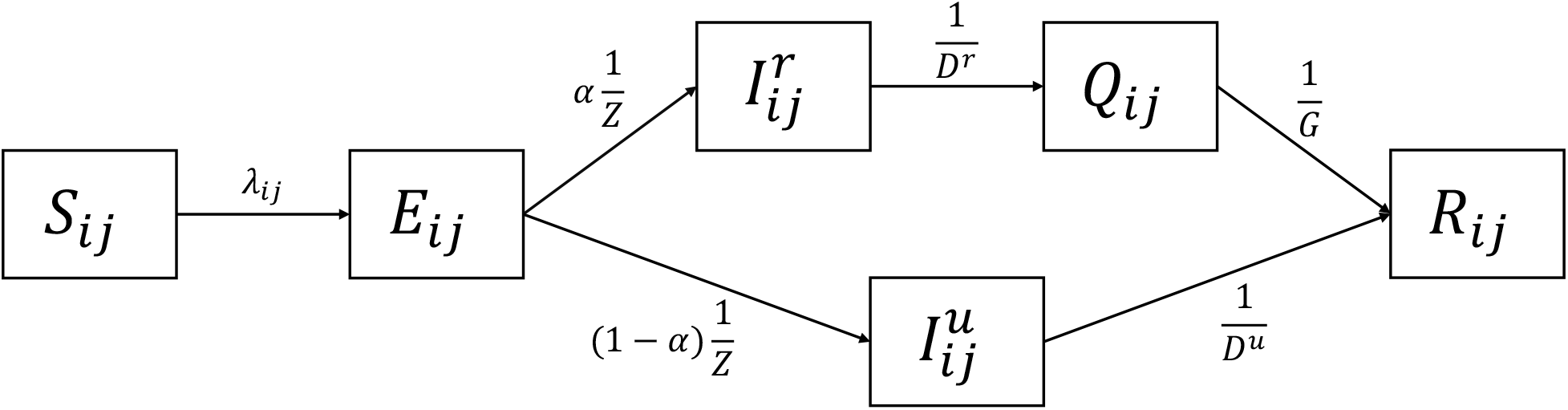
Transmission model diagram. *S*_*ij*_: susceptible, *E*_*ij*_: exposed, 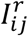 : reported infectious, 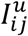: unreported infectious, *Q*_*ij*_: quarantined, and *R*_*ij*_: recovered. Parameters are described in Table 1.

At night, both 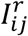 and 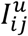 are assumed to stay within their residential areas, and the population size of region *j* is the sum of the number of people living in region *j* as shown in equation (9). Reported individuals 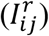 are infectious for *D*^*r*^ days on average until testing and quarantine, whereas unreported individuals 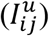 are infectious for *D*^*u*^ days on average and then recovered or removed (*R*_*ij*_). All reported cases 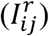 are quarantined (*Q*_*ij*_) immediately after being tested, reflecting South Korea’s measure, where self-quarantine was advised until receiving test results. After an average *G*-day quarantine, individuals recover or are removed (*R*_*ij*_). Given South Korea’s centralized public health governance, we assumed parameters μ, *α, D*^*r*^, *D*^*u*^, and *G* have the same value across the entire region. We also assumed no individuals enter or leave the model and no immunity loss following the primary infection as the simulation time period is relatively short.

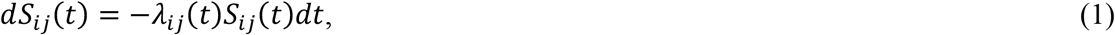

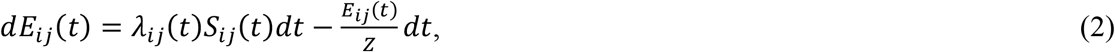

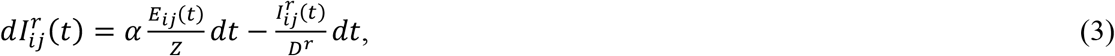

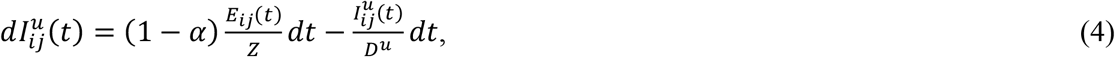

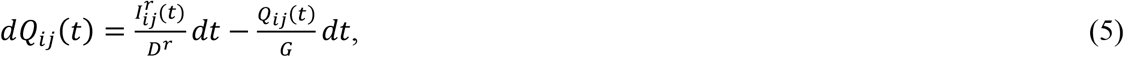

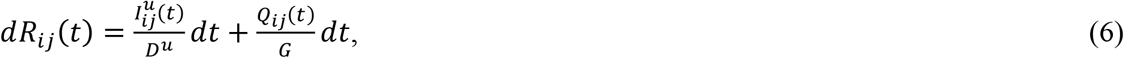

where the force of infection

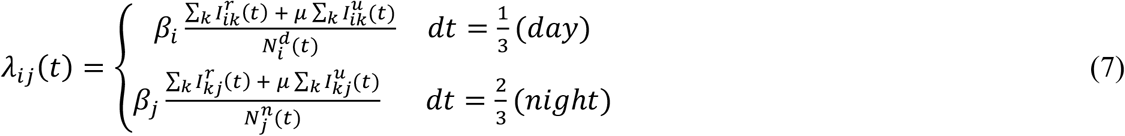

and

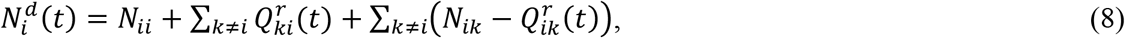

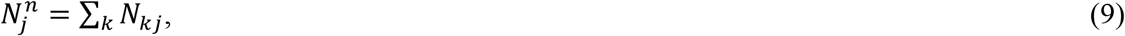

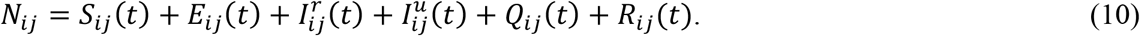

To impose stochasticity on the model, we assumed each term on the right hand side of equations (1)—(6) is a random variable (*u*_*i*_) that follows a Poisson distribution with a mean value described in equations (11)—(16). We then updated the state variables, as shown in equations (17)—(22), by randomly drawing a positive integer (*u*_*i*_) at each time step.

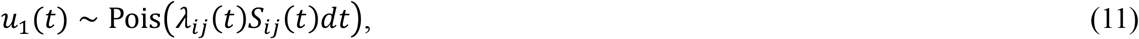

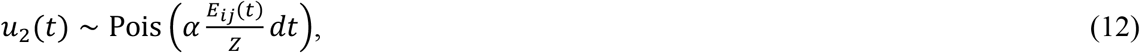

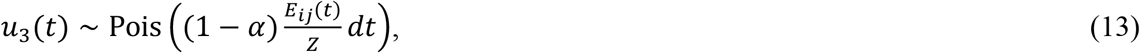

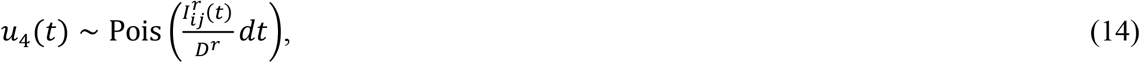

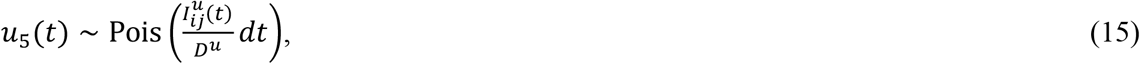

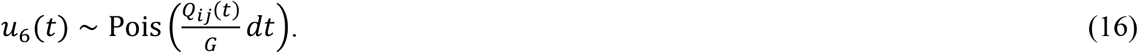

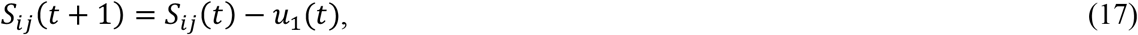

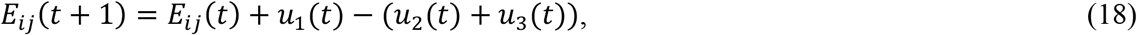

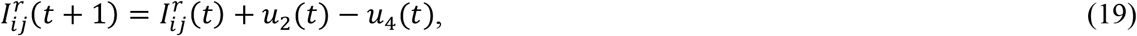

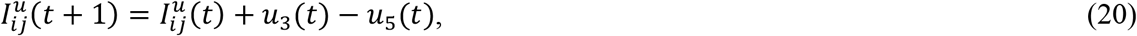

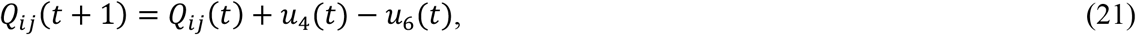

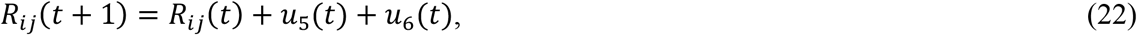

### Model calibration

The Ensemble Adjustment Kalman Filter (EAKF) infers the state of a dynamic system by assimilating a series of observations in conjunction with integration of an ensemble of simulations with varying initial conditions. Initially developed for numerical weather prediction, the EAKF has been adopted for use with infectious disease forecasting and analysis, including influenza models [26] and, more recently, COVID-19 models [16, 17, 25].

To estimate the distribution of the system state at time *t* given observations up to the current time *t*(**Y**_*t*_), the Kalman filter approach uses Bayes’ rule, as shown in equation (23), and assumes normality of the likelihood and prior distribution, allowing the posterior distribution to be completely characterized by the mean and covariance.

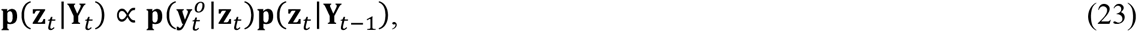

where 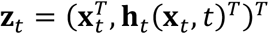 is the joint state-observation vector, combining the state vector (**x**_*t*_) and the expectation of the observation vector (**h**_*t*_ (**x**_*t*_,*t*)), **Y**_*t*_ is the set of all observations that are taken at or before time *t*, and 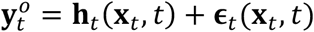 is the observation vector taken at time *t*, and **ϵ**_*t*_ (**x**_*t*_, *t*) is the error vector at time *t*.

In the EAKF, the prior and posterior mean and covariance are computed from the ensemble mean and covariance. Given observations from multiple locations, the filter processes these data sequentially to avoid more expensive computation [27]. Detailed computation of the posterior ensemble is presented in Algorithm 1 in the Supplementary Information.

### Filter divergence

A common challenge in the application of filtering methods is filter divergence, a situation in which the filter estimates deviate gradually from the truth over time. This divergence occurs when the ensemble variance of the model shrinks over successive assimilation of observations to values much less than the observational error variance (σ^*y*^)^5^. As a consequence, the filter gain becomes small, and new observations have a minimal impact on the observed state estimate [28]. To counter potential divergence, we applied multiplicative inflation [29, 30], as described in equation (24). To further improve the inference, we also utilized the space re-probing technique that randomly replaces a small proportion of ensemble members [31].

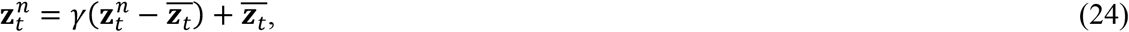

where 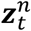 is the *n*th ensemble member of ***z***_*t*_, γ is the multiplicative inflation factor, and 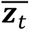 is the mean vector of ***z***_*t*_ across the ensemble.

### Model initialization

The size of each subpopulation was derived from 2020 census data from the KOSTAT [21] and 2019 mobility data from the KTDB [22]. The census data provided the population size of each si/do, which we denote as *P*_*j*_ (*j* = 1, 2, …, 17). The mobility data were assembled into a matrix, *M*_*ij*_ (*i, j* = 1, 2, …, 17), where *i* represents the destination and *j* represents the origin location. We initialized the off-diagonal subpopulations (*N*_*ij*_, *i* ≠ *j*) with *M*_*ij*_ and the diagonal subpopulations (*N*_*jj*_) with *P*_*j*_ − ∑ _*i*≠ *j*_ *M*_*ij*_.

Changes in the mobility matrix over time were also considered using regional population movement data updated weekly by the KOSTAT [23]. We first computed the relative change in the movement of the current week compared to the prior week from the data and multiplied it by the mobility matrix (*M*_*ij*_). We then reinitialized each subpopulation *Nij* according to the newly updated mobility matrix as described above, while preserving the proportions of each epidemiological stage.

The first wave in South Korea began with a sudden surge of case numbers after the identification of the 31^st^ case in Daegu on February 18, 2020 [24]. This patient was reported to have had symptoms and visited a medical facility, church, and hotel several days before being confirmed [32, 33]. Given these circumstances, it is presumed that multiple people had already been infected prior to February 18^th^, and one study estimated that 4 people (95% CI: 2—11) were already infectious but undocumented in Daegu on February 7^th^ [34]. Based on these reports, we began our simulation on February 7^th^ 2020 and initialized *E*_*ii*_ and 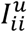 by randomly sampling from a uniform distribution with limits between 0 and 5. The initial value of *E*_*ij*_ and 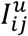 for all other subpopulations (*i* ≠ *j*), 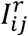, and *Q*_*ij*_ were set to zero. For *S*_*ij*_, we sampled from a uniform distribution, where the lower limit is 90% of the population (*N*_*ij*_), and the upper limit is *N*_*ij*_ − sum of upper limits of 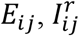 and 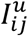 to have 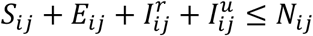. Lastly, *R*_*ij*_ was initialized by subtracting 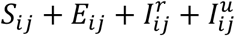 from *N*_*ij*_, so that 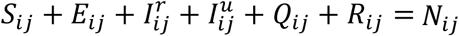.

The initial prior ranges for model parameters were derived from previous COVID-19 studies. For each parameter we sampled ensemble members from a uniform distribution with the initial prior ranges shown in Table 1.

**Table 1.** Model parameters with descriptions, initial prior ranges, limits, and references.

| Parameter | Description | Initial prior range | Limits | References |
| --- | --- | --- | --- | --- |
| $\beta_i$ | Transmission rate of region $i$<br>($i = 1, 2, \dots, 17$). | 0.3—1.6 | 0—5 | [9, 10, 16, 35] |
| $\mu$ | Relative infectivity of<br>unreported infections. | 0.1—0.7 | 0—1 | [16, 35] |
| $Z$ | Average latency period | 2—5 days | 1—6 days | [9, 16, 35, 36] |
| $\alpha$ | Ascertainment rate | 0.1—0.75 | 0—1 | [16, 17, 25, 35] |
| $D^r$ | Average infectious period of<br>reported cases | 1—3 days | 0.01—4 days | [16, 35, 36] |
| $D^u$ | Average infectious period of<br>unreported cases | 2—6 days | 1—7 days | [16, 35, 36] |
| $G$ | Average quarantine period | 12—15 days | 10—17 days | [36] |

We initialized 300 ensemble members per simulation and carried out ten independent simulations to accommodate the stochasticity inherent in the initialization process. The dynamics of the state variables evolved based on the model equations, and the EAKF updated all variables including model parameters within the predefined ranges outlined in Table 1.

### Observational data

We utilized the daily number of confirmed cases reported by the KDCA as observations. Specifically, we acquired the data at the si/do level for all 17 regions in South Korea, from February 8, 2020 to February 25, 2021. This period covers three waves of COVID-19 prior to vaccine introduction. These daily time series data exhibited significant noise and variability, potentially due to factors like reporting delays, testing availability, and policy changes. Using such data in the EAKF process produces spiky model fitting that captures short-term fluctuations rather than underlying trends. To reduce such overfitting, we smoothed the data by applying a 7-day moving average and used the smoothed time series data as γ^*o*^. The corresponding observed variable in the model was computed by integrating 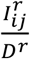 in equation (3) over time, equivalent to *u*4 in equation (19), and aggregated by residence to obtain the daily incidence for each of the 17 regions. The observational error variance 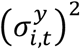 was set to 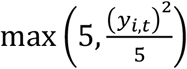, following a structure used in previous studies [16, 17, 25].

### Parameter estimation

We estimated *β* _*i*_ separately for each region and *α* at a nationwide level while keeping the remaining parameters, μ, *Z, D*^*r*^, *D*^*u*^, and *G* constant. For each parameter, we sampled ensemble members from a uniform distribution of the initial prior ranges specified in Table 1. By including *β* _*i*_ and *α* in the EAKF process and excluding the other parameters, we let *β* _*i*_ and *α* move within the boundary specified in Table 1 while the other parameters retain their initial values over time. A daily 1% multiplicative inflation (γ =1.01 in equation (24)) was applied to the observations and *β* _*i*_, and a weekly 1% re-probing was implemented for *β* _*i*_ and *α*. If a variable moved outside the range of its prescribed limits during the EAKF, inflation, or re-probing, its value was resampled to remain within the permissible range.

### Basic reproduction number

The basic reproduction number (*R*_0_) is defined as the number of secondary infections in a completely susceptible population caused by a single infected individual. Here, we approximated *R*_0_ as *βi*(*αD*^*r*^ + (1 − *α*)*μD*^*u*^), computed using the next generation matrix approach [37] for a model that assumes no movement between locations. The full model, equations (1)—(6), coupled with EAKF, produces time-varying parameter estimates. We thus defined two hyperparameters: 1) the time-varying basic reproduction number, *R*_*t*_ = *β* _*i,t*_ {*αtD*^*r*^ + (1 – *α*_*t*_)*μD*^*u*^}; and 2) the time-varying effective reproductive number,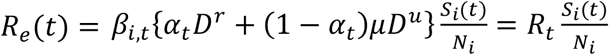. The key difference between *R*_*t*_ and *R*_*e*_ is that *R*_*t*_ assumes a fully susceptible population, whereas *R*_*e*_ reflects a partially susceptible population. Initially, these two measures start at the same value, but over time, they diverge as the number of infected people increase and the susceptible population is depleted. Here, we primarily focused on estimating *R*_*e*_ over the course of multiple COVID-19 waves in South Korea.

### Synthetic test

To verify the identifiability of the model parameters, we generated synthetic outbreaks using the SEIQR model run with prescribed parameter combinations and then conducted tests to determine if the full SEIQR-EAKF system could accurately identify those parameter combinations from the synthetic observations. Concretely, we created 100 combinations of parameters (*β* _*i*_, μ, *Z, α, D*^*r*^, *D*^*u*^, and *G*), by randomly drawing values from a uniform distribution of the given initial prior ranges (Table 1) and then used these parameters in free simulation to generate 100 different synthetic outbreaks. We estimated *β* _*i*_ and *α* and fixed the remaining parameters. To examine the overall convergence of the parameters we measured the error between the truth and the mean posterior estimate at the end of the outbreak. After analyzing the parameter identifiability with the synthetic data, we applied the identical inference framework and settings to the real data, aiming to estimate the identifiable parameters.

## Results

South Korea experienced three waves of COVID-19 prior to broad administration of vaccines, as shown in Figure 2. To explore estimates at specific time points, we picked dates before and after each wave. The first wave in South Korea hit Daegu on February 18, 2020, and spread to the adjacent area, North Gyeongsang. This outbreak lasted until the spring season and subsided by May 5, 2020 (date 1). Afterwards, sporadic outbreaks occurred across the region for approximately three months. On August 3, 2020 (date 2), the second wave began in Seoul and spread to the neighboring region of Gyeonggi. The number of cases in both regions returned to a low level by October 11, 2020 (date 3); however, the third wave arrived shortly thereafter. This wave affected all regions and continued until February 25, 2021 (date 4), immediately before vaccinations began.

**Figure 2.**
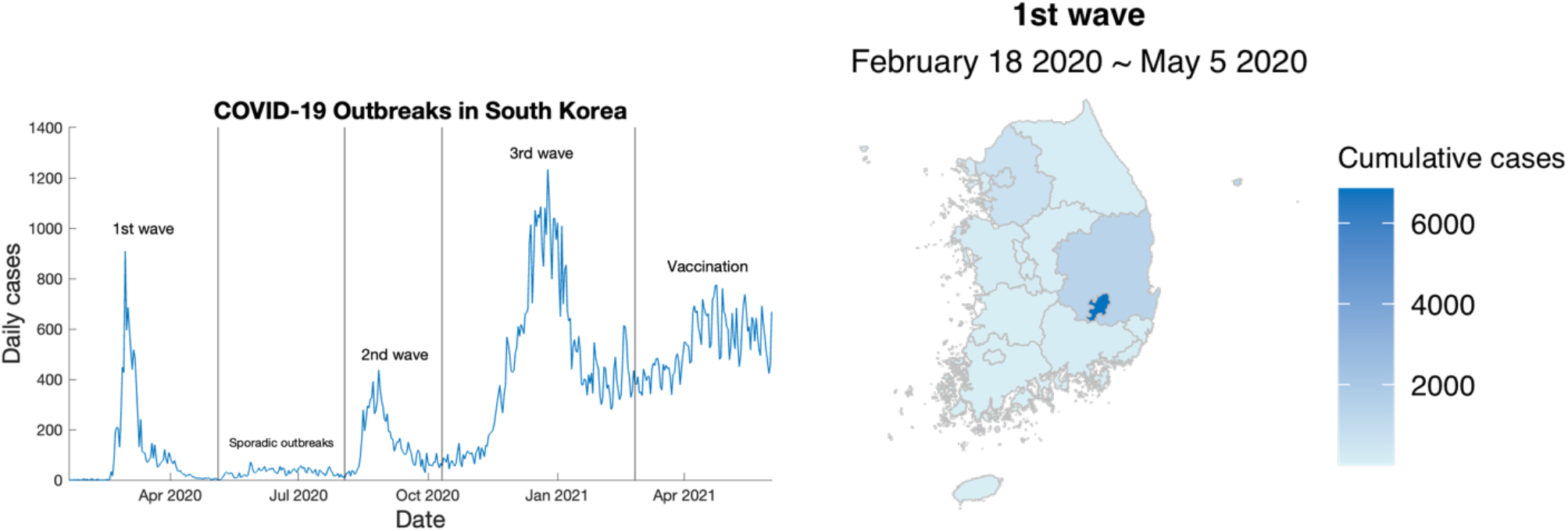
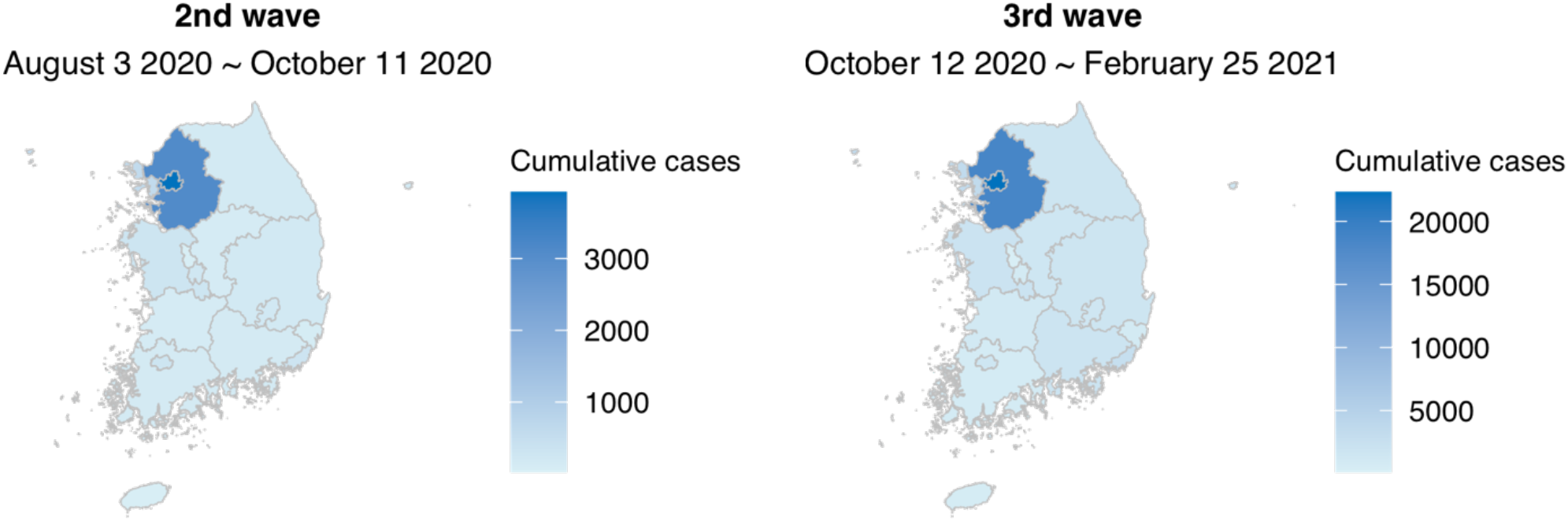
COVID-19 outbreaks in South Korea until February 25, 2021. The time series graph shows the total number of daily cases in South Korea and the vertical black lines represent the four selected dates, May 5, August 3, October 11, 2020, and February 25, 2021. Three geographic heat maps represent cumulative cases during each wave.

The estimates and error distributions for *β* _*i*_and *α* obtained from synthetic data are presented in the Supplementary Information (Figure S1). The posterior distribution of the model fitted to actual smoothed si/do data captures observed case levels across all regions. The mean closely aligned with the smoothed data and the 95% credible interval captured the unsmoothed original data (Figure 3, Figure S2).

**Figure 3.**
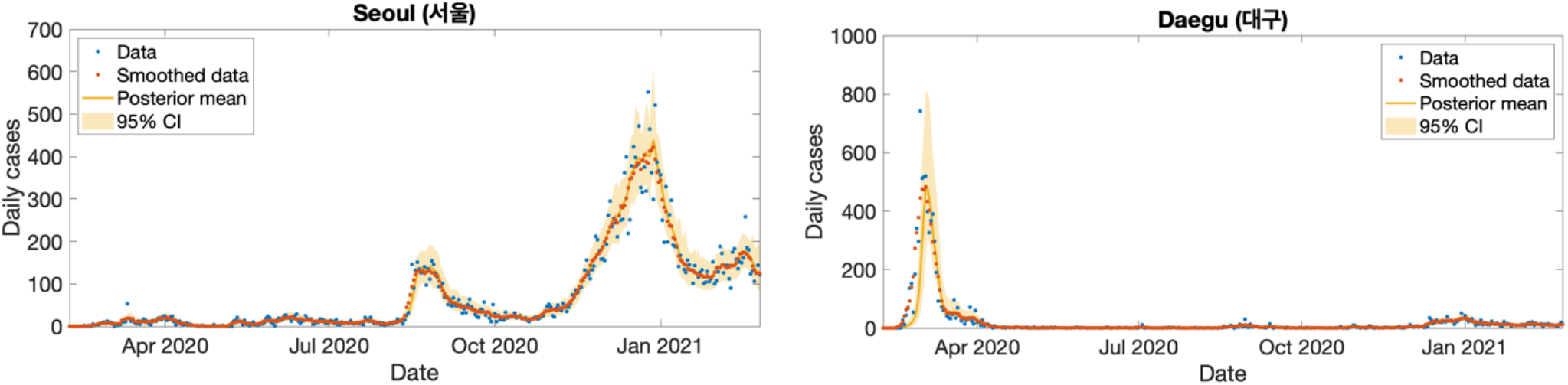
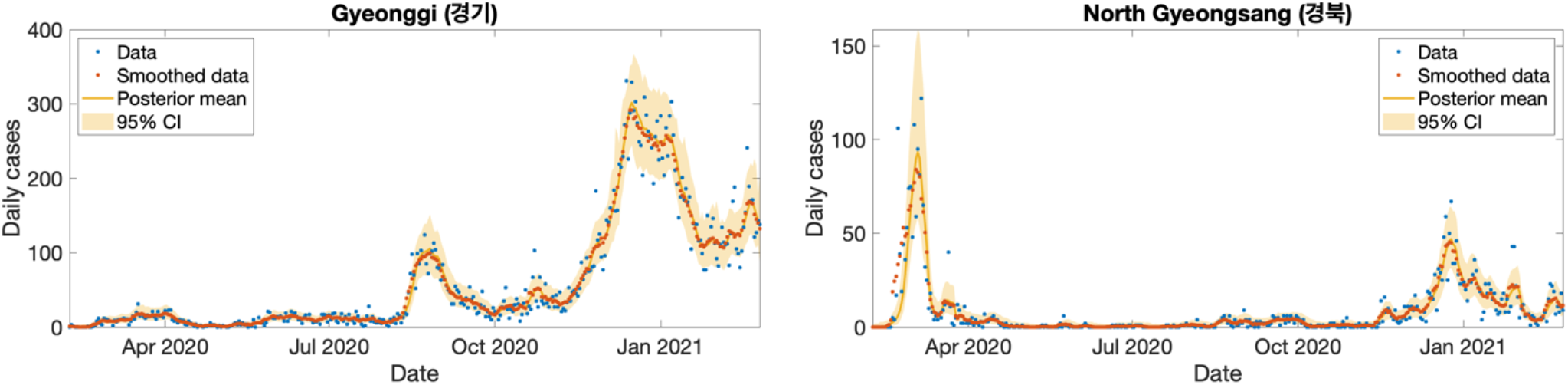
Daily confirmed cases data and posterior estimates for Seoul, Daegu, Gyeonggi, and North Gyeongsang from February 8, 2020 to February 25, 2021. The blue dots represent daily confirmed cases reported by the Korea Disease Control and Prevention Agency, the orange dots represent these data smoothed using a 7-day moving average, the yellow line and shaded area represent the posterior mean and 95% credible interval. Model fit for the remaining 13 regions are provided in the Supplementary Information (Figure S2).

During the entire simulation period, the model estimated similar transmission rates (*β* _*i*_) for Seoul and Gyeonggi due to their geographical proximity and substantial volume of inter-regional movement (Figure 4, Table 2). Meanwhile, the transmission rate estimates between Daegu and North Gyeongsang exhibited a little more difference with greater uncertainty in North Gyeongsang. The nationwide ascertainment rate (*α*) gradually increased over time (Figure 5, Table 2). After the first wave alone, the posterior mean estimate of the nationwide *α* reached 0.5 and over the course of the subsequent waves it increased to 0.62 due to the widespread testing and contact tracing in South Korea. The mean effective reproduction numbers (*R*_*e*_) of Daegu decreased and remained below 1 after the first wave (Figure 4). In the North Gyeongsang region, adjacent to Daegu, the mean *R*_*e*_ estimate showed a similar pattern. In the Seoul and Gyeonggi regions, the mean estimate of *R*_*e*_ fluctuated around 1 during sporadic outbreaks (May 3 – August 2, 2020). As the second wave began *R*_*e*_ increased to 2.14 in Seoul and 1.75 in Gyeonggi in mid-August but decreased to near 1 in early October. The third wave began in Seoul and spread nationwide, but the mean *R*_*e*_ in all regions hovered around 1.

**Table 2.**
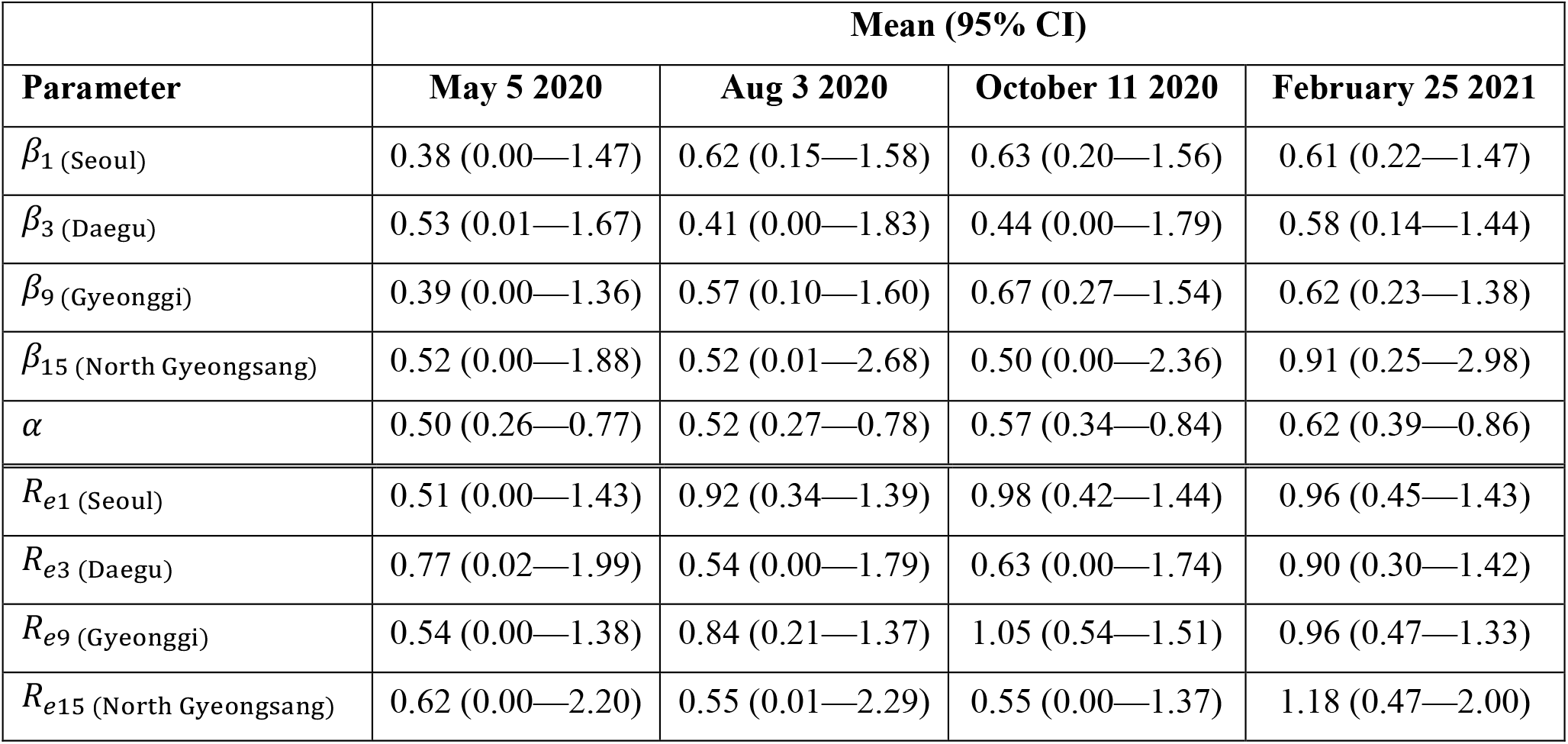
The mean and 95% credible interval (CI) of *β* _*i*_, *α*, and *R*_*e*_.

| Parameter | Mean (95% CI) |  |  |  |
| --- | --- | --- | --- | --- |
|  | May 5 2020 | Aug 3 2020 | October 11 2020 | February 25 2021 |
| $\beta_1$ (Seoul) | 0.38 (0.00—1.47) | 0.62 (0.15—1.58) | 0.63 (0.20—1.56) | 0.61 (0.22—1.47) |
| $\beta_3$ (Daegu) | 0.53 (0.01—1.67) | 0.41 (0.00—1.83) | 0.44 (0.00—1.79) | 0.58 (0.14—1.44) |
| $\beta_9$ (Gyeonggi) | 0.39 (0.00—1.36) | 0.57 (0.10—1.60) | 0.67 (0.27—1.54) | 0.62 (0.23—1.38) |
| $\beta_{15}$ (North Gyeongsang) | 0.52 (0.00—1.88) | 0.52 (0.01—2.68) | 0.50 (0.00—2.36) | 0.91 (0.25—2.98) |
| $\alpha$ | 0.50 (0.26—0.77) | 0.52 (0.27—0.78) | 0.57 (0.34—0.84) | 0.62 (0.39—0.86) |
| $R_{e1}$ (Seoul) | 0.51 (0.00—1.43) | 0.92 (0.34—1.39) | 0.98 (0.42—1.44) | 0.96 (0.45—1.43) |
| $R_{e3}$ (Daegu) | 0.77 (0.02—1.99) | 0.54 (0.00—1.79) | 0.63 (0.00—1.74) | 0.90 (0.30—1.42) |
| $R_{e9}$ (Gyeonggi) | 0.54 (0.00—1.38) | 0.84 (0.21—1.37) | 1.05 (0.54—1.51) | 0.96 (0.47—1.33) |
| $R_{e15}$ (North Gyeongsang) | 0.62 (0.00—2.20) | 0.55 (0.01—2.29) | 0.55 (0.00—1.37) | 1.18 (0.47—2.00) |

**Table 3.** Total cumulative number of infections by region as of February 25, 2021. Total infection includes both reported and unreported infections. Total infection rates = Total infections / Population × 100 (%). CI: credible interval.

| Region (Si/Do) | Population | Total infections (95% CI) | Total infection rates (95% CI) |
| --- | --- | --- | --- |
| Seoul | 9,586,195 | 57,651 (35,085—121,415) | 0.60 (0.37—1.27) |
| Busan | 3,349,016 | 6,833 (4,089—13,192) | 0.20 (0.12—0.39) |
| Daegu | 2,410,700 | 19,180 (8,977—51,679) | 0.80 (0.37—2.14) |
| Incheon | 2,945,454 | 9,291 (5,704—19,578) | 0.32 (0.19—0.66) |
| Gwangju | 1,477,573 | 4,219 (2,613—8,818) | 0.29 (0.18—0.60) |
| Daejeon | 1,488,435 | 2,601 (1,527—6,017) | 0.17 (0.10—0.40) |
| Ulsan | 1,135,423 | 2,138 (1,273—4,816) | 0.19 (0.11—0.42) |
| Sejong | 353,933 | 651 (294—2,065) | 0.18 (0.08—0.58) |
| Gyeonggi | 13,511,676 | 47,799 (29,398—91,507) | 0.35 (0.22—0.68) |
| Gangwon | 1,521,763 | 3,909 (2,439—9,097) | 0.26 (0.16—0.60) |
| North Chungcheong | 1,632,088 | 3,693 (2,234—8,365) | 0.23 (0.14—0.51) |
| South Chungcheong | 2,176,636 | 5,147 (3,174—10,380) | 0.24 (0.15—0.48) |
| North Jeolla | 1,802,766 | 2,484 (1,479—5,457) | 0.14 (0.08—0.30) |
| South Jeolla | 1,788,807 | 1,961 (1,123—4,601) | 0.11 (0.06—0.26) |
| North Gyeongsang | 2,644,757 | 7,244 (3,985—16,647) | 0.27 (0.15—0.63) |
| South Gyeongsang | 3,333,056 | 4,672 (2,864—9,744) | 0.14 (0.09—0.29) |
| Jeju | 670,858 | 1,313 (712—3,171) | 0.20 (0.11—0.47) |
| Nationwide | 51829023 | 180,788 (109,827—355,983) | 0.35 (0.21—0.69) |

**Figure 4.**
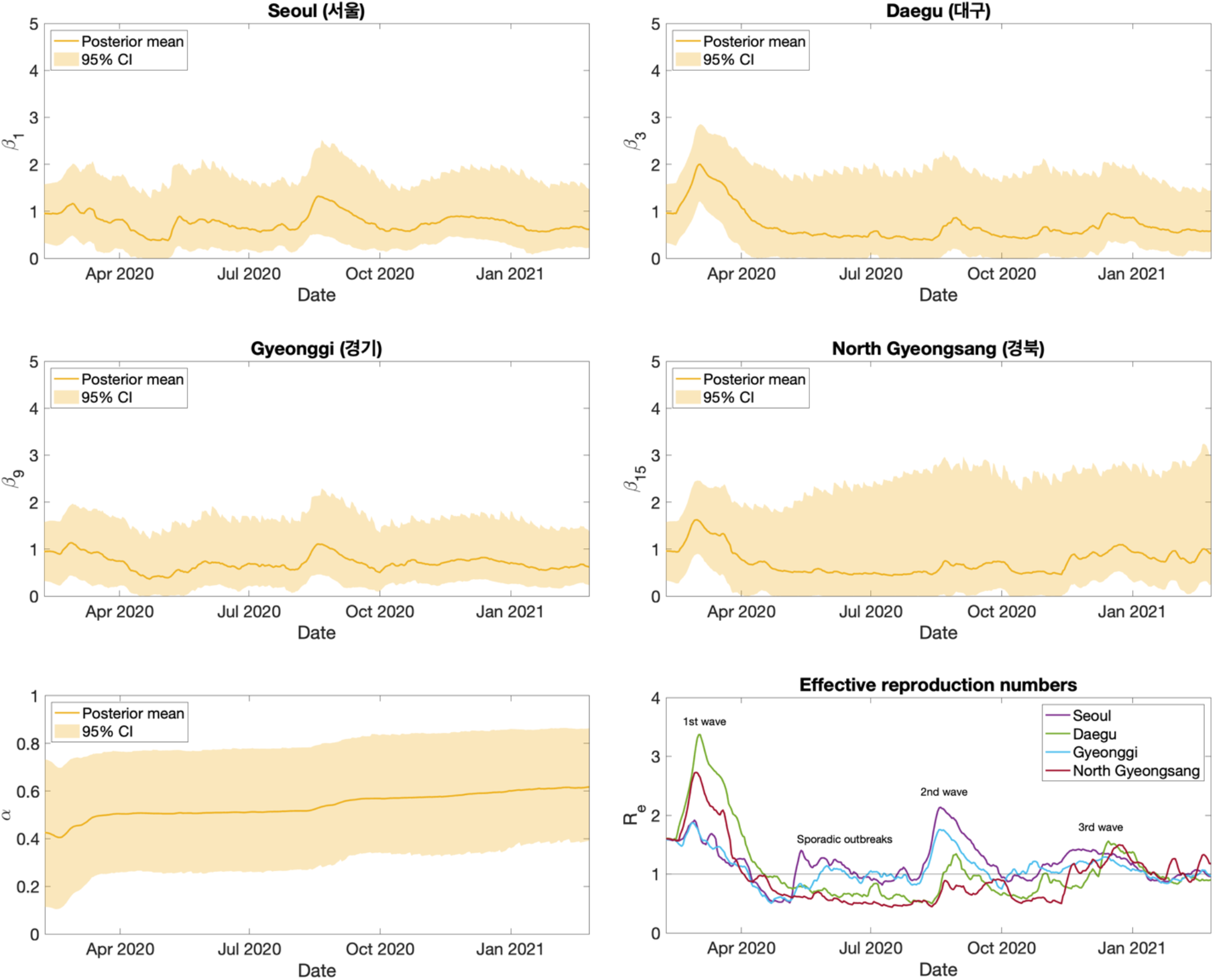
Posterior mean and credible intervals of parameters from February 8, 2020 to February 25, 2021. The posterior mean effective reproduction number for the Seoul, Daegu, Gyeonggi, and North Gyeongsang areas during the same time horizon. The transmission rates for the remaining 13 regions are provided in the Supplementary Information (Figure S3).

As of February 25, 2021, the total infection rates, including reported and unreported infections, in Seoul and Daegu were higher than in other regions, as these locations were the epicenters of major waves. The next highest total infection rates were estimated for the Gyeonggi and Incheon regions, which are included in the Seoul metropolitan area. Overall, the mean estimate of cumulative total infection rates was less than 1% across all regions, and the nationwide estimate was also less than 1%.

## Discussion

This study presents a metapopulation model combined with the EAKF and describes the characteristics of the first 13 months of the COVID-19 pandemic in South Korea. During that period, COVID-19 cases, hospitalizations, and deaths were fewer per capita than in most other countries. Simulations with the SEIQR-EAKF system were used to infer the transmission rate, ascertainment rate, and reproduction number over the courses of three waves and cumulative infections just before vaccination.

Our 95% credible interval estimates of the transmission rate in Daegu (0.01—1.67) and North Gyeongsang (0.00—1.88) on May 5, 2020 (Table 2), cover the findings from another study, which inferred country-level transmission rates of 0.162 and 0.449 for the periods February 29– March 13 and March 14–April 29, respectively [10]. That study utilized a SEIHR (Susceptible-Exposed-Infectious-Hospitalized-Recovered) model structure with the hospitalized compartment analogously functioning as the quarantine compartment in our model. However, the SEIHR model did not represent undocumented infections and simulated the entire Korean population, i.e. it had no spatial structure. Another study reported a higher transmission rate of 4.62 for Daegu and North Gyeongsang [13]. Similar to our approach, this study employed a SEIQR model structure, but introduced an additional susceptible population (*S*_*F*_) consisting of individuals whose behavior changed in response to disease control interventions. Individuals from the regular susceptible population (*S*) transitioned into the behavior-changed population (*S*_*F*_) at a fixed rate, and the transmission rate for this group was set at 2% of the rate for the regular susceptible population. These distinctions played a crucial role in driving the higher transmission rate.

The estimate of the ascertainment rate (*α*) suggests that around half of infections were tested and confirmed, whereas the other half were not documented, and that this ratio increased steadily across the first three waves of the pandemic in South Korea. This finding is in close alignment with a prior study that used a probabilistic model to quantify undetected COVID-19 cases in South Korea and which estimated between 10,400 and 139,900 undetected infections by February 2, 2021, leading to a total estimated infection range of 89,244 to 218,744 [18]. Dividing the cumulative number of confirmed cases as of February 2, 2021 (78,844) by the total cases (89,244—218,744) yields a range of 0.360—0.883, closely matching the 95% credible interval estimated here for *α* on February 25, 2021 (0.39—0.86) (Table 2). This contrasts significantly with the numbers from China and the United States, where the proportion of documented cases was estimated at 0.14 (0.10—0.18) before the initiation of travel restrictions on January 23, 2020 in China [16], and 0.245 (0.186—0.323) during December 2020 in the US [17]. South Korea’s success in identifying and containing a higher proportion of infected individuals throughout 2020 may have been facilitated by its immediate and comprehensive implementation of testing and screening protocols from the very beginning the pandemic.

The estimated total number of infections from our model largely corroborates, yet slightly exceeds, the previous study estimate of 89,244 to 218,744 [18]. Comparatively, South Korea’s cumulative total infection rate were markedly lower than those in many other countries. A study from the US estimated 69.0% (63.6—75.4%) population susceptibility at the end of 2020, implying that about 30% of population had been infected [17]. The reported cases alone in the US were more than 18.9 million, corresponding to about 5.7% of the population in 2020. In other countries, documented case rates by the end of 2020 were 2.0% in Germany, 3.4% in Italy, 3.5% in UK, 4.1% in Spain, 4.4% in Netherland, 2.5% in Türkiye, 2.1% Russia, 1.2% in Mexico, 3.5% in Brazil, and 3.6% in Argentina [38]. All these values exceed the estimated total infection rate (0.35%) for South Korea on February 25, 2021.

The mean estimate of effective reproduction number (*R*_*e*_) also clearly demonstrates the impact of South Korea’s efforts to contain COVID-19. Even before the initial surge in cases, South Korea had already increased diagnostic capacity by producing test kits and securing testing facilities [2]. Upon the arrival of the first wave they implemented meticulous contact tracing, using credit card transaction history and closed-circuit television footage. These measures coupled with social distancing and mask wearing, but without locking down cities or closing the borders, quickly reduced the reproduction number during the first and second waves, preventing the spread of infection nationwide, and kept the reproduction number around one after the second wave. Our findings corroborate the effectiveness of the Korean government’s systematic preparedness and rapid response to COVID-19.

In interpreting the findings of our study, it is crucial to recognize its limitations. The mobility trend data we employed to adjust the mobility matrix over time did not differentiate between intra- and inter-regional movements at the si/do level. As a consequence, we used aggregated data to adjust the subpopulations involved in inter-regional movements. In the model structure, loss of immunity and reinfection after primary infection were not explicitly included because the simulation time horizon was short. Another potential limitation is that delays in diagnosis and reporting were not considered in our analysis. In South Korea, the testing turnaround time and the time from testing to reporting were both less than a day [39-41], which was short relative to other countries. To address and minimize the impact of such delays, we smoothed the case data by averaging over 7 days, which enabled focus on underlying outbreak trends. Lastly, the synthetic data used to test the model-inference system lacked real-world complexities, such as control interventions that could alter parameter values during an outbreak. Nevertheless, employing synthetic data remains an effective strategy for understanding the identifiability of stochastic models such as the SEIQR used here.

In summary, our study provided insights into COVID-19 transmission dynamics in South Korea by estimating the transmission rate and ascertainment rate through the SEIQR-EAKF model framework. Our findings highlight the critical role that swift public health interventions, including extensive testing and proactive contact tracing, had in controlling the spread of the virus. As we navigate forward, these lessons reinforce the importance of pandemic preparedness and response efforts, offering a valuable reference point for other countries.

## Data Availability

All data and code used to produce the results and analyses presented in this manuscript are available from Github repository

https://github.com/creamy11/metapop_covid19_kor

