## Supplementary Information for "COVID-19 transmission dynamics in South Korea prior to vaccine distribution"

**Algorithm 1**. Sequential ensemble adjustment Kalman filter

---

**Input**: observations $\left\{\mathbf{y}_{t_k} = \left(y_{1,t_k}, y_{2,t_k}, \dots, y_{n\_loc,t_k}\right)^T\right\}_k$, observational error variance $\left\{\left(\sigma^y_{i,t_k}\right)^2\right\}_{i,k}$ for $i = 1,2,\dots,n\_loc$ and $k = 1,2,\dots,nt$.

**Initialize** $\mathbf{z}^n_{t_0} = \left(x^n_{1,t_0}, x^n_{2,t_0}, \dots, x^n_{n\_loc,t_0}, w^n_{t_0}, h^n_{1,t_0}, h^n_{2,t_0}, \dots, h^n_{n\_loc,t_0}\right)^T$ for $n = 1,2,\dots,n\_ens$.

**for** $k = 1$ to $nt$ **do**

  **for** $n = 1$ to $n\_ens$ **do**

    Solve the model with $\mathbf{z}^n_{t_{k-1}}$ and obtain $\mathbf{z}^n_{t_k}$

  **end for**

  Do inflation and re-probing.

  **for** $i = 1$ to $n\_loc$ **do**

    Compute $\left(\sigma^h_{i,t_k}\right)^2 = \mathrm{Var}\left(\left\{h^n_{i,t_k}\right\}_n\right)$

    **for** $n = 1$ to $n\_ens$ **do**

      Compute

$$\hat{h}_{i,t_k}^n = \frac{\left(\sigma_{i,t_k}^y\right)^2\left(\sigma_{i,t_k}^h\right)^2}{\left(\sigma_{i,t_k}^y\right)^2+\left(\sigma_{i,t_k}^h\right)^2}\left(\frac{\overline{h_{i,t_k}}}{\left(\sigma_{i,t_k}^h\right)^2} + \frac{y_{i,t_k}}{\left(\sigma_{i,t_k}^y\right)^2}\right) + \sqrt{\frac{\left(\sigma_{i,t_k}^y\right)^2}{\left(\sigma_{i,t_k}^y\right)^2+\left(\sigma_{i,t_k}^h\right)^2}}\left(h_{i,t_k}^n - \overline{h_{i,t_k}}\right)$$

Compute $dw_{i,t_k}^n = \frac{\text{Cov}\left\{w_{t_k}^n, h_{i,t_k}^n\right\}}{\left(\sigma_{i,t_k}^h\right)^2}\left(\hat{h}_{i,t_k}^n - h_{i,t_k}^n\right)$

Update $x_{i,t_k}^n = x_{i,t_k}^n + \frac{\text{Cov}\left\{x_{i,t_k}^n, h_{i,t_k}^n\right\}}{\left(\sigma_{i,t_k}^h\right)^2}\left(\hat{h}_{i,t_k}^n - h_{i,t_k}^n\right)$

Update $h_{i,t_k}^n = \hat{h}_{i,t_k}^n$

**end for**

**end for**

**for** $n = 1$ to $n\_ens$ **do**

Update $w_{t_k}^n = w_{t_k}^n + \frac{\sum_i dw_{i,t_k}^n}{n\_loc}$

**end for**

**end for**

**Output:** $\mathbf{z}_{t_k}^n = \left(x_{1,t_k}^n, x_{2,t_k}^n, \ldots, x_{n\_loc,t_k}^n, w_{t_k}^n, h_{1,t_k}^n, h_{2,t_k}^n, \ldots, h_{n\_loc,t_k}^n\right)^T$ for $k = 1, 2, \ldots, nt$ and $n = 1, 2, \ldots, n\_ens$.

---

At time $t_k$ and location $i$, $x_{i,t_k}^n$ denotes $n$th ensemble member of the unobservable local variable; $w_{t_k}^n$ denotes $n$th ensemble member of the unobservable global variable; $h_{i,t_k}^n$ denotes $n$th ensemble number of observable variable; $\left(\sigma_{i,t_k}^h\right)^2$ denotes ensemble variance; $\overline{h_{i,t_k}}$ denotes ensemble mean of $h_{i,t_k}^n$. For an unobserved variable we further updated the *n*th ensemble member by scaling the difference between prior and posterior of the observed variable using the covariability between the observed and unobserved variables across ensemble members and adding the scaled difference value to the prior. For the variables homogeneous throughout all regions ($w_{t_k}^n$), we updated them by averaging the Kalman gain computed from the observation in each region. In our model, $x_{i,t_k}^n$ corresponds to the state variables or $\beta_{i,t_k}$, and $w_{t_k}^n$ corresponds to $\alpha_{t_k}$.

*System identifiability*: Given the model structure and observations from one data set, $\beta_i$ and $\alpha$ converged well to the truth and were identifiable (Figure S1). The overall convergence of all 17 $\beta_i$ and $\alpha$ had an error distribution centering around zero (Figure S1).

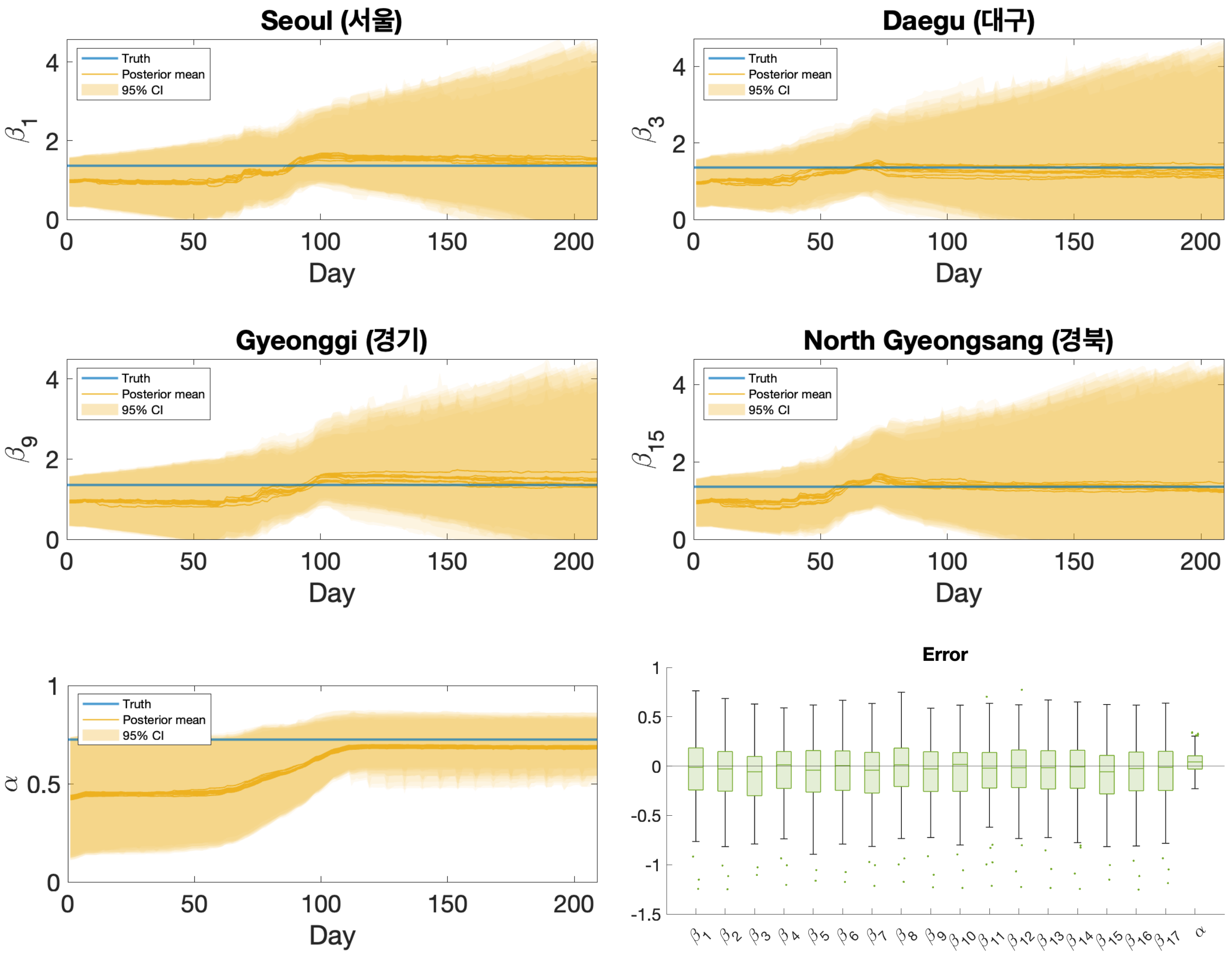


**Figure S1. Convergence of parameters in the synthetic data.** Five time series plots show the convergence of $\beta_i$ and $\alpha$ estimates from synthetic outbreak data after fixing parameters, $\mu$, $Z$, $D^r$, $D^u$, and $G$. In each plot, the blue line represents the truth, the yellow line and shaded area represent the mean and the 95% credible interval (CI) of the posterior distribution of ten independent simulations. The box chart shows errors between the posterior mean and truth aggregated from 100 different synthetic datasets. The errors are computed at the end of the outbreak.

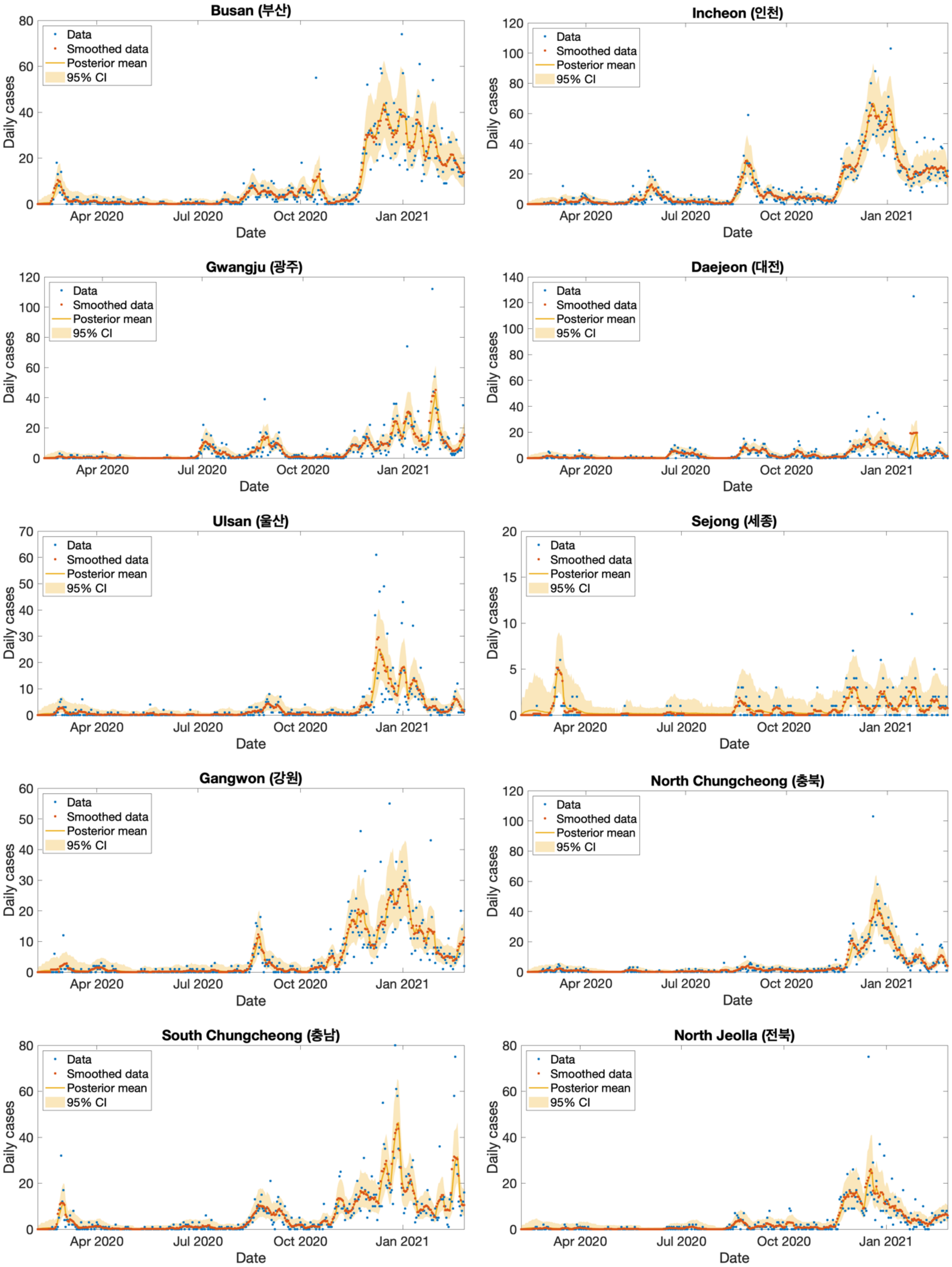
Busan (부산)
Incheon (인천)
Gwangju (광주)
Daejeon (대전)
Ulsan (울산)
Sejong (세종)
Gangwon (강원)
North Chungcheong (충북)
South Chungcheong (충남)
North Jeolla (전북)
Data
Smoothed data
Posterior mean
95% CI
Daily cases
Date
Apr 2020
Jul 2020
Oct 2020
Jan 2021

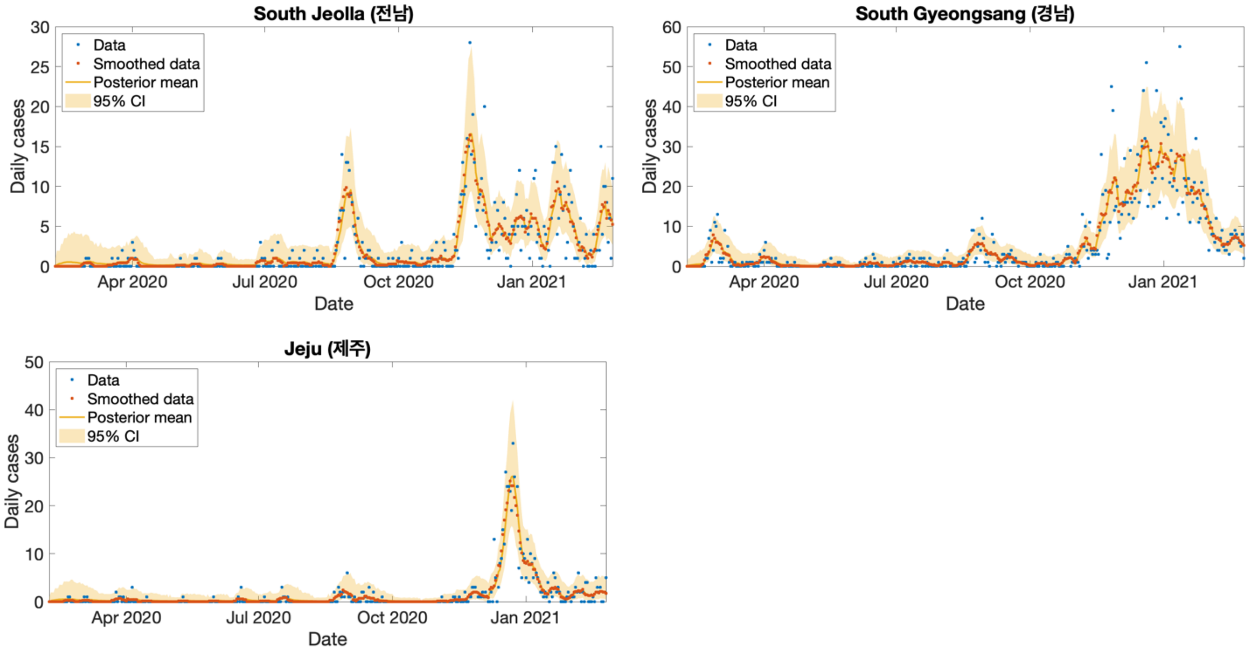


**Figure S2.** Daily confirmed cases data and the posterior estimates from February 8, 2020 to February 25, 2021. The blue dots represent daily confirmed cases reported by Korea Disease Control and Prevention Agency, the orange dots represent smoothed data by using a 7-day moving average, and the yellow line and shaded area represent the posterior mean and 95% credible interval.

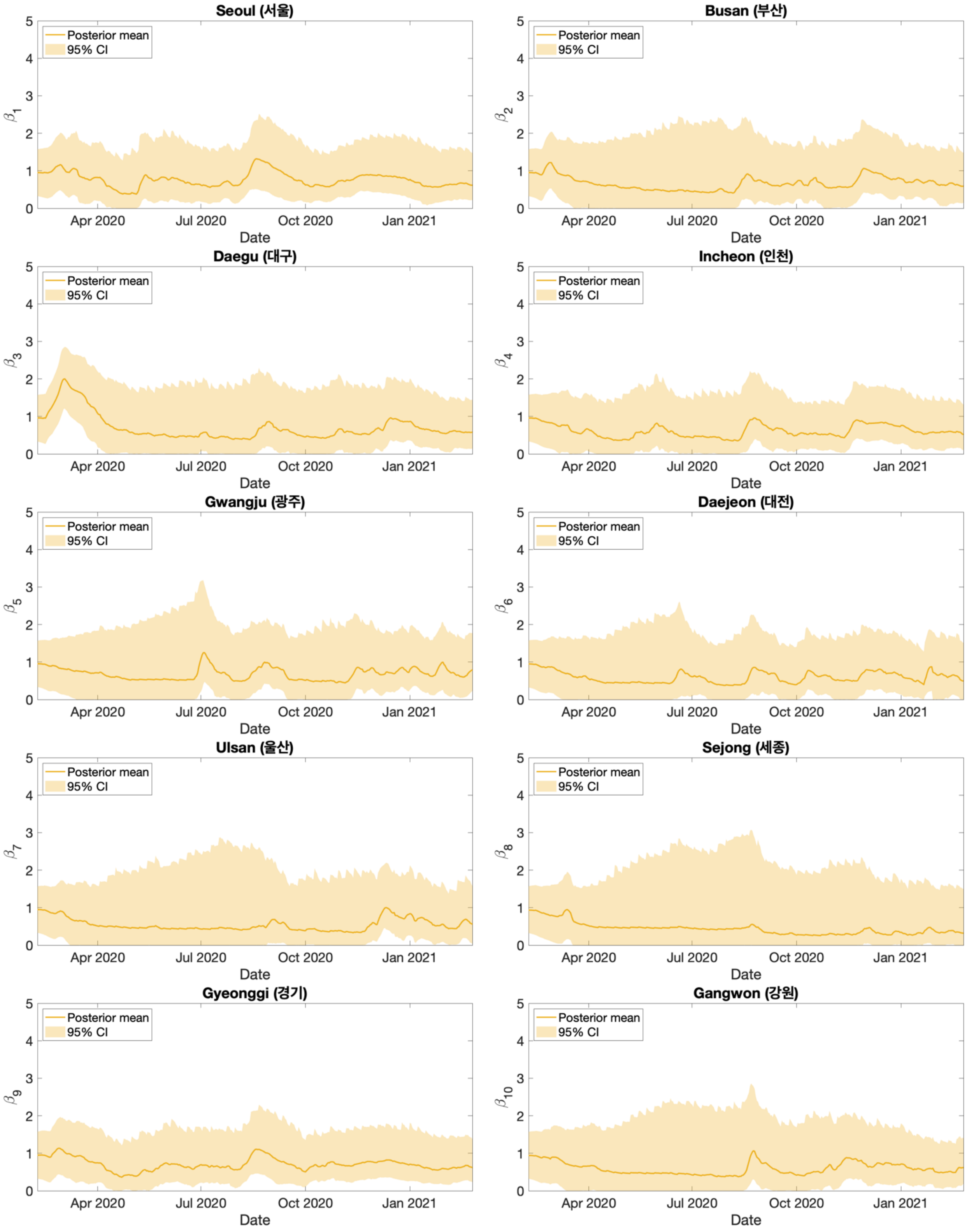
Seoul (서울)
Busan (부산)
Daegu (대구)
Incheon (인천)
Gwangju (광주)
Daejeon (대전)
Ulsan (울산)
Sejong (세종)
Gyeonggi (경기)
Gangwon (강원)
Posterior mean
95% CI
$\beta_1$
$\beta_2$
$\beta_3$
$\beta_4$
$\beta_5$
$\beta_6$
$\beta_7$
$\beta_8$
$\beta_9$
$\beta_{10}$
0
1
2
3
4
5
Apr 2020
Jul 2020
Oct 2020
Jan 2021
Date

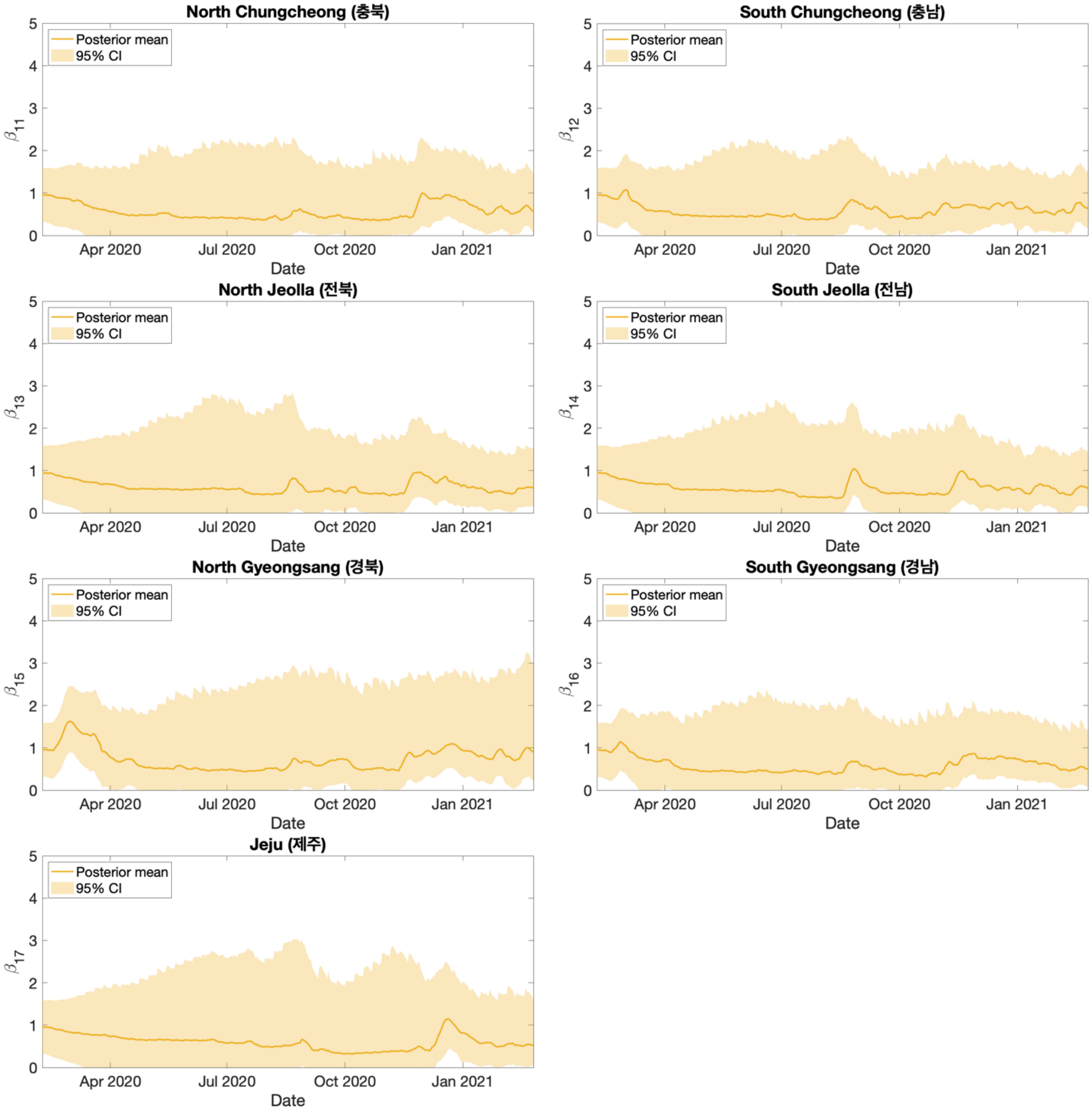


**Figure S3.** Posterior mean and 95% credible intervals of the transmission rate from February 8 2020 to February 25 2021.
